# Language, perceived discrimination and barriers to complaint use: sub-Saharan African immigrants’ healthcare experiences in Oslo, a qualitative interview study

**DOI:** 10.64898/2026.09.07.26361905

**Authors:** Peter Kofi Taadi, Bill Derman

**Affiliations:** Institute for Health Policy and Research (IHPR), Kumasi, Ghana; Department of International Environment and Development Studies (Noragric), Faculty of Landscape and Society, Norwegian University of Life Sciences (NMBU), Ås, Norway

**Keywords:** Emigrants and immigrants, health services accessibility, communication barriers, perceived discrimination, right to health, healthcare candidacy, accountability, qualitative research, Norway

## Abstract

Universal coverage alone does not ensure equitable healthcare. Racism is consistently associated with poorer healthcare experiences, but not consistently with lower healthcare use. What patients do after an encounter they interpret as discriminatory therefore remains unclear. This study examined how sub-Saharan African immigrants in Oslo interpreted such encounters, how language shaped comprehension and participation, how those experiences related to later service use, and what their accounts indicated about rights and accountability. We interviewed 15 first-generation immigrants from five sub-Saharan African countries, individually and in English, in 2021. Recruitment was purposive and referral-based. We analysed the data using reflexive thematic analysis. A working definition of discrimination was provided before participants described events. Awareness of the Equality and Anti-Discrimination Ombud and Tribunal was prompted rather than spontaneously recalled. We applied candidacy and a right-to-health framework, covering availability, accessibility, acceptability, quality, participation and accountability, abductively after initial theme development. Ten participants reported encounters they interpreted as discriminatory, including dismissive conduct, an explicitly group-based remark, ambiguous double-gloving and inadequate language support. All five contrasting accounts included communication described as adequate in a shared language, which we advance as a tentative linguistic recognition hypothesis rather than as evidence of cause. Inadequate communication affected reported comprehension and participation, and troubling encounters were followed by continued attendance, avoidance or self-medication. When prompted, all participants agreed that discrimination violated human rights. None described making a formal complaint, and their accounts point to five overlapping mechanisms of non-use: anticipated futility, emotional conservation, normalisation, procedural uncertainty and relational risk. Attendance can therefore record clinical need or absent alternatives rather than acceptable care. Healthcare use is an inadequate equity indicator. Complaint mechanisms that go unused generate institutional records showing no problem. Integrating candidacy, a right-to-health framework and the naming, blaming and claiming sequence, the study proposes language accessibility as a testable determinant of recognition, participation and accountability. It does not establish legal violations or causal effects.

## Introduction

Norway finances a largely tax-funded health service and grants residents statutory patient rights, a registered general practitioner and a statutory right to interpretation. At the start of 2026, immigrants comprised 17.5% of the population and Norwegian-born people with immigrant parents a further 4.2% [1]. Formal entitlement of this kind is often treated as settling the question of access. It does not. Access is a process rather than a status. It involves whether services can be reached, understood, trusted and used safely, and whether an encounter leaves a patient willing to return. A system can extend the same rights to everyone and still deliver unequal care. This study is situated in that gap between entitlement and encounter.

We define discrimination as unjustified different treatment or disadvantage linked to protected or racialised characteristics [2]. Two related terms serve different purposes here. ’Perceived discrimination’ refers to participants’ own interpretations of an encounter, whilst ’racialisation’ refers to our analysis of how differences in ethnicity, nationality, language or skin colour acquired social meaning. Perceived discrimination matters even where intent or legal violation cannot be established, because patients act on how they understood an encounter and on what they expect next time.

Racism is a public health concern in two connected ways. It is associated with poorer mental and physical health [3,4]. It also shapes the conditions of access itself: communication, treatment decisions, dignity and trust. Both patterns are well documented in European healthcare. Norwegian studies report language difficulties, stereotyping and divergent interpretations of what equal access means among sub-Saharan African users and healthcare workers [5,6], patterns of distrust and “othering” among immigrant patients in Norwegian hospitals [7], and racism named explicitly as a help-seeking barrier by East African migrants [8]. Work with Syrian refugees in Norway locates the same dynamics inside the clinical encounter itself, where the user’s perception of the caregiver, communication and time determine whether the care journey builds trust or distrust [9]. Nationally representative survey evidence links perceived discrimination to substantially higher odds of mental health problems, moderated by belonging and trust [10]. European scoping reviews converge on the same interpersonal and structural mechanisms, and identify limited translation, prejudice and not being taken seriously as the recurring content of these encounters [11–15]. They also converge on the same limitation. Evidence remains concentrated in a small number of national settings and in refugees and recent migrants rather than racialised populations more broadly [12,16].

Where the evidence stops converging is on what any of this does to healthcare use. Racism is consistently associated with poorer experiences, weaker trust and worse communication, but not consistently with lower utilisation [17]. That inconsistency is usually read as a measurement problem. It may instead be substantive. Continued attendance can reflect clinical need or the absence of alternatives rather than acceptable care. Utilisation counts may therefore record persistence whilst concealing harm. UK meta-ethnographic work makes the opposite pattern visible in mental healthcare, where fear of harm and prior racist treatment produce avoidance and disengagement [18]. What separates a patient who keeps attending from one who withdraws is not established. Nor is it known what the first group carries into later encounters.

This study therefore asks what patients do after an encounter they interpret as discriminatory. Continued use, avoidance and informal self-treatment are different outcomes with different consequences. Only the first is visible in routine data. We use candidacy theory because it treats access as jointly and continually negotiated between people and services rather than as a fixed entitlement [19]. This allows a discriminatory encounter to be located at a specific stage of that negotiation rather than treated as a diffuse atmosphere. Where perceived exclusion converts into avoidance or self-medication, an equity problem becomes a public health risk.

A second question concerns what happens after that. Health and equality systems in Norway provide routes for advice, review and redress. The existence of a mechanism does not establish that the people it was built for can find it, understand it, or use it without risk to their continuing care. Research elsewhere shows healthcare organisations absorbing discrimination complaints into general patient-experience processes, whilst potential complainants anticipate individualisation, dismissal or emotional cost [20–22]. This matters beyond individual redress. A complaint system that racialised patients do not use produces institutional records showing no problem. The most recent European review of inpatient racism reaches this point directly, closing on the need for robust, transparent and accessible complaint procedures [12]. These interviews were not designed as a complaint-behaviour study, and we make no claim to measure complaint rates. What participants volunteered about rights, institutions and non-use is analysed as evidence of how accountability appeared from where they stood.

Norway is an analytically instructive setting for both questions. Its formal protections against discrimination are unusually extensive, whilst routine measurement of racialised inequality remains limited. Where the formal side is close to maximal, the distance between entitlement and lived accessibility is easier to see. This assumes nothing about whether the Norwegian system is uniquely inclusive or exclusionary. It also addresses the setting concentration those reviews identify, since Norway is among the European contexts they name as under-represented [12,16]. The section that follows sets out the relevant legal and institutional arrangements, and the conceptual framework after it brings three lenses to bear: candidacy on how access is negotiated, a right-to-health framework on the conditions that make access equitable, and the naming, blaming and claiming sequence on whether an interpreted wrong becomes a claim. Integrating the three is this study’s principal analytic contribution.

The study asked (1) how participants described and interpreted healthcare encounters they considered discriminatory or unequal; (2) how language and interpreting affected comprehension, participation and perceived dignity; (3) how these encounters related to continued healthcare use, avoidance or informal care; and (4) how participants’ accounts indicated awareness of rights and access to complaint and accountability mechanisms. The study does not determine legal violations. It examines how universal rights on paper were experienced through the practical conditions of access.

Establishing what patients do after a discriminatory encounter changes what the inconsistent utilisation evidence can be taken to mean. It separates a system that retains racialised patients from one that merely records them. For services, it identifies the stage at which an encounter turns a patient away, which is where an intervention has to sit. For equality institutions and health authorities in Norway and comparable systems, it indicates whether accountability mechanisms reach the patients they were built for. Their silence may not mean that nothing is wrong. Read together, the three frameworks also offer an account of how language accessibility, the negotiation of candidacy and the mobilisation of rights connect, which each on its own leaves open.

### Legal and institutional context

The International Convention on the Elimination of All Forms of Racial Discrimination is incorporated through Section 5 of the Equality and Anti-Discrimination Act and applies as Norwegian law [23]. It is not incorporated into the Human Rights Act. It therefore does not carry the same precedence rule as conventions incorporated there [24]. This distinction is institutional.

It should not be confused with an absence of legal protection against racial discrimination. A court resolving a conflict between ICERD-derived rights and an ordinary statute does not apply the automatic priority that Human Rights Act conventions receive. ICERD nonetheless remains binding domestic law under the Equality and Anti-Discrimination Act.

The Equality and Anti-Discrimination Act prohibits direct and indirect discrimination on grounds including ethnicity, religion and belief [23]. For the Act, ethnicity includes national origin, descent, skin colour and language. ’Race’ is not enumerated as a separate protected ground. The stated legislative concern was that the terminology might reinforce biological notions of separate human races [24]. The UN Working Group of Experts on People of African Descent [25] however argued that the omission may constrain race-conscious public discourse. The relevant point here is narrower than legal ’unnameability’. Norwegian law protects characteristics commonly implicated in racial discrimination, but organises them under ethnicity. Language and skin colour therefore sit within the same statutory category. This does not establish that every language barrier constitutes ethnic or racial discrimination. It does mean that linguistic disadvantage can be legally relevant where it produces unjustified differential effects associated with ethnicity. This study did not assess whether the reported encounters met that legal threshold.

Statistics Norway does not routinely produce register statistics based on self-identified ethnic background. Population statistics use country of birth, parental country background and immigration category [24]. These variables permit important analyses. They do not directly measure skin colour, racialisation or self-identified ethnicity. The resulting gap constrains population-level monitoring of racialised inequalities in healthcare rather than making it impossible [25,26].

The Equality and Anti-Discrimination Ombud offers free advice and can guide individuals considering a complaint. The Anti-Discrimination Tribunal determines complaints and may issue binding decisions and, in qualifying cases, compensation. Healthcare users may also complain to the provider, request a supervisory case from the County Governor, submit a rights appeal, seek assistance from the Health and Social Services Ombudsman, or claim compensation for patient injury [27]. Government figures show relatively few Tribunal complaints registered on ethnicity grounds between 2018 and 2022, most concerning working life [24]. Those figures indicate low formal use. They do not by themselves establish institutional futility, nor show that no healthcare-related cases occurred. This study examines what participants said about these institutions and what they associated with not pursuing formal redress.

The Interpreting Act entered into force on 1 January 2022, after the fieldwork. It requires public bodies to use an interpreter where necessary to uphold legal safeguards or provide proper services, and it sets requirements for interpreter qualifications [28]. In healthcare, the need for an interpreter depends on whether patient and professional can communicate adequately. It cannot be inferred from immigrant status alone. National patient guidance places responsibility for assessing that need on the healthcare professional, responsibility for booking a qualified interpreter on the service, and rules out children and unqualified relatives in that role [29]. Earlier research in Norway however documented ’invisible rights’. Polish migrants described receiving information they did not fully understand, and staff who were reluctant to book interpreters or overestimated their Norwegian proficiency [30].

These arrangements are what make Norway analytically useful. Protection is extensive on paper, so where language needs, racialisation and uncertainty about complaint procedures still overlap in practice, the gap lies between design and experience rather than between design and absence. They do not determine how any individual encounter should be classified.

### Conceptual framework

Utilisation is an incomplete measure of equitable access. Candidacy conceptualises eligibility for healthcare as jointly and continually negotiated between people and services [19]. Relevant dimensions include recognition of need, navigation, service ease of entering and using services, presentation for care, professional response and local operating conditions [19,31]. In this study, linguistic work bears on ease of entering and using services, while interpersonal conduct bears on perceived professional response. Continued attendance represents continued pursuit of care. Avoidance or self-medication represents reduced engagement with it. Candidacy is used interpretively, not as evidence that reported conduct caused later behaviour.

A public-health analysis of access must therefore ask more than whether people attend. It must ask whether communication, dignity, trust and available alternatives make attendance meaningful and safe. The framework allows continued use to coexist analytically with dissatisfaction, and positions avoidance and informal treatment as potential consequences rather than individual failures.

The human right to the highest attainable standard of health includes timely, appropriate and good-quality healthcare without discrimination. General Comment No. 14 identifies availability, accessibility, acceptability and quality as interrelated elements of that right. Contemporary WHO and OHCHR guidance also emphasises participation and accountability [32–34]. Accessibility includes informational and linguistic access, as well as non-discrimination. Acceptability concerns dignity, medical ethics and respect for persons, and quality includes clinically adequate communication. Participation requires sufficient information and communicative support for patients to understand, question and contribute to decisions.

These standards are used as an evaluative public-health framework rather than a judicial test. The interviews cannot determine whether an encounter was unlawful discrimination or a breach of an international obligation. They can identify circumstances that suggest possible concerns about equitable accessibility, acceptability, quality or participation, and examine how patients interpreted those circumstances. Racialisation describes how ethnic, national, linguistic or skin-colour differences acquired social meaning in participants’ accounts. Perceived discrimination denotes participants’ own interpretations.

Accountability is part of the right to health, because rights require mechanisms through which concerns can be raised, reviewed and remedied. Legal-consciousness scholarship distinguishes recognising an injury, attributing responsibility and making a claim [35,36]. Accountability cannot however be reduced to individual willingness to complain. Meaningful mechanisms must be visible, understandable, independent, accessible and safe for people who may need continuing care.

Non-discrimination rights may be difficult to mobilise [37], and institutions may reclassify rights claims as service-quality matters [20]. People may anticipate disbelief, futility, bureaucracy or emotional cost [22,38]. Complaint capability is therefore treated as a relational and sensitising concept. It concerns the interaction between a potential complainant’s knowledge and resources and the accessibility and credibility of institutional mechanisms. The interviews did not systematically measure procedural knowledge or complaint use.

The analytic sequence links encounter, interpretation, candidacy response, service-use response and possible use of accountability. It is not a causal model, and not every participant moved through every stage. Its purpose is to connect public-health consequences with the rights-based conditions of accessibility, participation and accountability. Candidacy concerns the negotiation of a claim to healthcare. Naming, blaming and claiming concerns whether an interpreted wrong can become a claim about the conditions of that care [35]. The two processes intersect, because a person weighing whether to complain may still depend on the institution being challenged. That is why this study treats complaint capability as distinct from candidacy, though shaped by it. Figure 1 summarises the integrated pathway.

**Figure 1.**
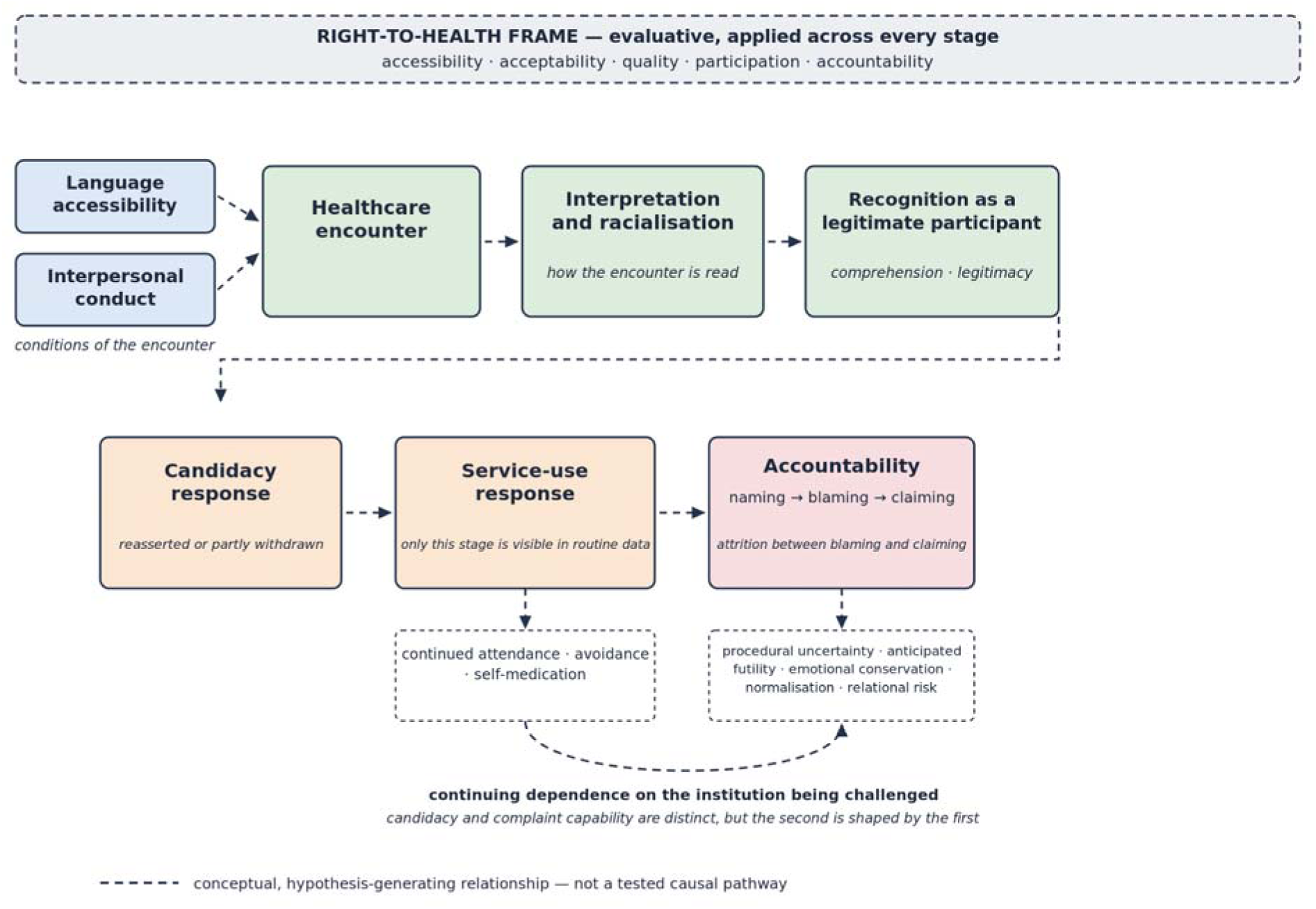
Integrated conceptual pathway linking language accessibility, candidacy and complaint capability within the right-to-health framework. Dashed arrows denote conceptual, hypothesis-generating relationships rather than tested causal effects. Not every participant moved through every stage.

## Methods

### Design overview

This exploratory qualitative interview study collected contextualised accounts of healthcare experiences. Reporting was informed by the Consolidated Criteria for Reporting Qualitative Research (COREQ) [39]. The completed checklist, coding tree and interview guide are provided as supplementary files.

### Participants and sampling

Participants were first-generation immigrants born in an African country and resident in Oslo. We used purposive sampling, followed by participant referral. Fifteen adults from five countries took part (Table 1): ten women and five men; four from Ghana, three each from Nigeria, Cameroon and Uganda, and two from The Gambia. Ages ranged from 25 to 45 years and length of residence from three to sixteen years. All had used Norwegian healthcare at least twice and had a regular general practitioner. The sample was relatively highly educated and cannot represent the diversity of African-origin residents. The original protocol excluded first language, preferred healthcare language and Norwegian proficiency. This was a scope decision made before fieldwork, not a later oversight. No directly approached eligible participant was recorded as declining, although informal non-response within referral chains could not be measured. We did not cap referral chains per seed participant or deliberately diversify across networks. That may have concentrated the sample within a small number of social circles and contributed to its educational skew.

**Table 1.** Participant characteristics.

| Pseudonym | Country | Sex | Years in Norway | Education | Migration route | Services used | Discrimination |
| --- | --- | --- | --- | --- | --- | --- | --- |
| Ali | Ghana | F | 4 | Senior secondary | Family reunion | GP, hospital, ED | No |
| Ama | Ghana | F | 16 | Bachelor | Family reunion | GP, hospital | Yes |
| Adwoa | Ghana | F | 5 | Vocational | Family reunion | GP, hospital | No |
| Kwasi | Ghana | M | 3 | Master | Studies | GP, hospital, ED | Yes |
| Oju | Nigeria | F | 3 | Bachelor | Family reunion | GP | Yes |
| Ada | Nigeria | F | 6 | Master | Family reunion | GP, hospital, ED | Yes |
| Ebe Ano | Nigeria | F | 4 | Master | Studies | GP, hospital | Yes |
| Divine | Cameroon | M | 3 | Master | Studies | GP | Yes |
| Philip | Cameroon | M | 5 | Master | Studies | GP, hospital, ED | Yes |
| Ogah | Cameroon | M | 4 | Senior secondary | Family reunion | GP, hospital, ED | No |
| Rose | Uganda | F | 1–5* | Master | Studies | GP | Yes |
| Juliet | Uganda | F | 15 | Basic education | Family reunion | GP, hospital, ED | No |
| Rob | Uganda | M | 10 | Bachelor | Work | GP, hospital, ED | Yes |
| Yube | Gambia | F | 4 | Senior secondary | Family reunion | GP, hospital, ED | No |
| Ogene | Gambia | F | 5 | Bachelor | Family reunion | GP, hospital, ED | Yes |

*Pseudonyms are used in the text. GP, general practitioner (fastlege); ED, emergency*
*department. Yes/No indicates whether the participant reported an encounter they interpreted as racial discrimination. First language, preferred language for healthcare and Norwegian proficiency were not recorded systematically and are therefore not tabulated; this bounds the shared-language comparison discussed in the limitations. \*Age and years in Norway are reported as bands rather than exact values for this row to reduce re-identification risk, as the data-protection note in the Methods explains.*

### Data collection

We conducted individual telephone interviews in English following ethical approval 19^th^ June 2020, during pandemic-related restrictions in Oslo. Participant recruitment and data collection took place from 10/08/2020 to 15/12/2020. Interviews lasted about one hour and were conducted privately, with no third party present. They were audio-recorded with consent and transcribed verbatim. We made brief field notes after each interview to record contextual observations. No interview was repeated. We deleted the recordings after analysis to prevent third-party access. The pilot-tested schedule covered background, healthcare encounters, possible discrimination, later service use, coping, and awareness of rights and institutions. We used two versions, branching on whether the participant reported experiencing discrimination. Both included the same rights and institutions section. The schedule is provided as supplementary material. Three questions covered the rights domain. Participants were asked whether they understood discrimination to be a violation of human rights. They were asked whether they knew of any international or non-governmental organisations advocating against discrimination in Norway, and could name them. They were also asked whether they knew of the Norwegian Equality and Anti-Discrimination institution, its appointed Ombud, and the Tribunal through which redress may be sought.

Three features of that design bound what the findings can support, and they are carried through to the reporting. The third question names the Ombud and the Tribunal, so awareness of those two bodies cannot be assumed to be unprompted. No question asked about complaint routes specific to health services, such as a provider’s complaints office, the Health and Social Services Ombudsman or the County Governor. The absence of reference to them therefore reflects the schedule as much as the participants. No question asked whether a participant had ever approached anyone. Accounts of non-use arose when participants elaborated on the awareness questions, rather than in response to a direct question about use.

Before participants recounted their experiences, the interviewer explained a working definition of racial discrimination. This ordering may have primed how participants classified ambiguous events. The analysis therefore distinguishes concrete descriptions, participants’ own attributions and the authors’ subsequent theoretical interpretation. The interview guide is provided as supplementary material.

The initial screening question distinguished the two branches of the schedule, and therefore the ten- and five-participant groups analysed in the results. It used a compound formulation. Participants were asked whether they had felt or experienced some form of “discrimination/racism/prejudice/unequal treatment” in Norwegian healthcare. Discrimination, racism, prejudice and unequal treatment are not conceptually equivalent. A question bundling all four is a looser elicitation instrument than one addressing discrimination alone. Some participants coded into the discrimination-reporting group may therefore be responding to perceived unequal treatment or prejudice more broadly. This affects the two-group classification used throughout the Results and Discussion. It is treated as a limitation of the elicitation method rather than of the participants’ accounts.

### Data analysis

Transcripts were analysed manually, without qualitative data analysis software, using reflexive thematic analysis [40,41]. Initial coding focused on reported events, meanings attributed by participants, language and interpretation, later service use, and knowledge or use of complaint procedures. The first author conducted the coding and discussed theme development with the second author and supervisor. Consistent with reflexive thematic analysis, that dialogue aimed to interpret and challenge themes rather than calculate inter-coder reliability. An audit trail, reflexive notes and attention to negative cases supported analytic transparency. Transcripts and findings were not returned to participants for comment or correction. The supplementary coding tree provides the final theme structure and representative codes.

The five mechanisms of complaint non-use require a note on their origin. Three of them, anticipated futility, emotional conservation and normalisation, correspond to codes generated during initial coding of the rights theme. Two others were assembled after coding. Relational risk draws on material coded under coping, where concern about jeopardising future care was expressed. Procedural uncertainty summarises the absence, across accounts, of any reference to a specific procedure. These two are therefore a re-description across themes rather than sub-themes produced by the coding, and the supplementary coding tree marks the distinction. We initially developed themes from the dataset. We then applied candidacy and the right-to-health framework abductively, to examine how themes related to service ease of entering and using services, dignity, participation, continuity and accountability. This sequence distinguishes participants’ accounts from later interpretation. The pre-interview definition of racial discrimination nonetheless means data generation was not wholly inductive. We analysed the five participants reporting no discrimination as contrasting cases. Theme development proceeded through repeated engagement with the complete dataset. We did not use saturation as a criterion for sample sufficiency, because the sample was fixed in advance.

Adequacy was judged by depth rather than by a target count set in advance. Interviews lasted about one hour and generated accounts detailed enough to address the encounter-classification, language and service-use questions for both groups. The sample is comparable in size to related qualitative studies of migrant healthcare experiences in Norway. These range from six interviews in Pettersen and Debesay’s study of East African migrants [8] to forty-seven in Mbanya et al.’s study of sub-Saharan African immigrants [5]. The five-participant contrasting group is correspondingly the most under-powered part of the analysis, as is any comparison by country or migration route within either group. Comparisons of that kind are not attempted, and observations that draw on the contrast are flagged as hypothesis-generating wherever they appear.

### Research team, reflexivity and ethics

The Norwegian Centre for Research Data (NSD; now Sikt) approved the study (reference no. 186966; 19 June 2020). Participants gave verbal informed consent for telephone interviewing, and pseudonyms are used throughout. The first author conducted the interviews as part of his Master of Science degree in International Relations at the Norwegian University of Life Sciences, supervised by the second author. He was then an international student living in Norway, with a professional background in public health and training in qualitative interviewing. His insider position supported access and rapport. It also created a risk of shared assumptions and over-identification, and reflexive notes and supervisory discussion helped examine these influences. The second author brought an external disciplinary vantage point. He is a social anthropologist whose career spans Michigan State University and the Norwegian University of Life Sciences (Noragric), and whose research on land, water and human-rights claiming in Africa informed his role in supervision, including challenging interpretations during theme development. Participants were recruited through community networks by referral. They were informed of the study’s aims and the researcher’s insider background before consenting. No participant had a prior personal or professional relationship with the interviewer beyond this referral connection.

Table 1 cross-tabulates pseudonym against country, sex, age, years in Norway, education, migration route, services used and discrimination status, for a small and identifiable community. Each row was therefore reviewed for uniqueness within the sample before submission. One row was judged to carry a compounded risk. The participant who described an informal, non-clinical channel for obtaining medicines is identifiable by a narrow combination of exact demographic values, alongside a description of an act that could carry legal or immigration consequences if she were recognised. For that row only, Table 1 reports age and years in Norway as five-year bands rather than exact values, and the quotation no longer names a specific country of origin. No other row met this compounded threshold, and no other data were altered.

## Results

### Interpersonal conduct and perceived classification

Ten participants reported at least one encounter they interpreted as racial discrimination; five did not. The themes that follow preserve the distinction between reported conduct, participants’ interpretation and the authors’ conceptual reading. Counts describe this sample only and are not prevalence estimates.

Participants most often described non-verbal conduct that they interpreted as signalling they did not belong, including facial expressions and body language. Telephone interviews and retrospective recall prevented independent assessment. The analysis therefore treats these as reported perceptions rather than observable classifications. Participants familiar with clinical work sometimes used professional expectations to explain their interpretation:

> *“The way she looked disgusted at me upon seeing me was very bad. It was as if I was some piece of trash. I am a nurse, and I know how healthcare workers welcome patients, with a handshake if possible, with smiles, and by making patients feel at ease. This is exactly the opposite of that.” (Ama)*

One participant described a healthcare worker putting on two pairs of gloves before venepuncture. He interpreted this as exceptional distancing, and worried that staff believed he was infectious. The study cannot establish whether double-gloving differed from the worker’s usual practice, reflected an infection-control habit, or had another clinical explanation. This episode is therefore presented as an ambiguous reported event, and as the meaning the participant attributed to it, rather than as proven differential treatment:

> *“The healthcare worker wore two sets of gloves right in front of me to take my blood sample. This was not the first time I had my sample taken. I became worried that perhaps I have a contagious infection which my doctor did not mention to me.” (Kwasi)*

The five participants assigned to the non-reporting branch following the compound screening question shared one feature: communication in a language shared with the provider, either Norwegian or English. Participants also described these encounters as cordial, respectful and competent. These cases are not a controlled comparison. They were not matched with the reporting group on language proficiency, length of residence, education or service type, and encounter complexity was not standardised, so what can be inferred from the contrast is limited.

### Language, interpreting and perceived exclusion

Language was central to many accounts. Participants described services defaulting to Norwegian, ad hoc use of English, and interpreters not being proactively offered. The study cannot determine whether an interpreter was clinically required in every episode, since adequacy depends on the complexity and stakes of communication. Participants’ accounts however show that language accessibility affected comprehension, participation and perceived legitimacy. Some explicitly interpreted limited accommodation as conditional belonging. Where they did not, the authors later interpreted it as racialisation. Several also read the absence of accommodation as placing responsibility for adapting to the encounter on them rather than on the service providing care.

> *“It was my first time giving birth at the hospital, and everything written and spoken was in Norwegian. The midwives tried to explain procedures in English rather than through a certified interpreter, which I did not really understand, but I was in so much pain that I just said yes to whatever they said.” (Ada)*

This account raises a public-health and rights concern about meaningful participation. The available data do not establish whether legal consent requirements were breached. They do indicate reported assent without adequate understanding, during a clinically vulnerable encounter. That engages informational accessibility, communication quality, autonomy and dignity, and it is consistent with broader evidence that racialised migrant women’s maternal-care encounters are often shaped by inadequate communication and reduced participation in decision-making [42]. National guidance advises against replacing qualified interpreters with unqualified relatives [29].

### Subsequent service use: continued attendance, avoidance and self-medication

Participants described different responses after troubling encounters. Several continued attending because they expected to need services for themselves or relatives. In some accounts, family responsibility was offered as one reason for preserving a workable relationship with providers. Philip described doing so after a doctor allegedly denied sick leave while stating that Africans were strong and did not fall sick:

> *“I was not happy he denied me sick leave, saying Africans are strong and therefore do not fall sick. We both laughed about it, but I was not happy within. I wanted to keep a good relationship because of the family and so ignored it.” (Philip)*

The quoted remark, if recalled accurately, is more explicit than the ambiguous non-verbal and double-gloving episodes. It assigns a group-based characteristic directly to the participant. Philip’s account also shows that continued attendance did not imply satisfaction. He described it as a pragmatic response to anticipated future need.

A smaller number of participants described delaying care, avoiding services or self-medicating. Rose, who lived alone, described abandoning the health service after repeated difficulties with language and obtaining medicines through an informal channel instead:

> *“I don’t even waste my time going to the health facility after I realised it was frustrating every time with language issues. I arranged for a friend to bring me common medications such as painkillers and antibiotics from abroad. I manage myself anytime I feel I am not well.” (Rose)*

Family composition and temporal sequences were not systematically measured. These accounts therefore do not establish that absence of family responsibility caused withdrawal. Withdrawal from mainstream services following experiences interpreted as racist has been documented elsewhere among minoritised users able to disengage [18]. Avoidance and self-medication are also recognised among the coping repertoires described by migrants who report discrimination [43].

Avoidance and silence also appeared in coping accounts. Most participants described weighing confrontation against a perceived risk to future care, and several linked that calculation to the absence of any route for raising a concern:

> *“For fear of being denied proper treatment, I just keep quiet. I am angry within, but what can I do? There is no channel to communicate your feelings and concerns apart from a service-rating text message one receives after leaving.” (Ebe Ano)*

Only one participant, a nurse, reported directly challenging a provider. These accounts establish participants’ concerns and choices, not that retaliation occurred. The family-obligation interpretation is therefore retained only as a provisional explanation for some cases, not as a cross-sample finding.

### Rights awareness and barriers to complaint use

When asked whether discrimination was a human-rights violation, all participants agreed. The question supplied the proposition, so this represents prompted agreement rather than spontaneous demonstration of legal knowledge. Some participants however questioned whether protections were practically available to them:

> *“In theory, I know discrimination or racism is a violation of human rights, but I think this violation is not for everyone. What rights do we have? None I practice. They know blacks do not have the confidence to speak up even when they are discriminated against.” (Ada)*

Asked whether they knew of organisations advocating against discrimination in Norway, participants named integration and immigration authorities, labour unions, human-rights bodies and the United Nations. Several of these are not complaint bodies at all, and none is specific to health services. The Equality and Anti-Discrimination Ombud and the Anti-Discrimination Tribunal were also discussed. A later question in the schedule named both, so participants cannot be reported as unaware of them.

No participant described approaching any of these bodies about the encounters they recounted. Participants volunteered this non-use in follow-up discussion rather than in answer to a direct question. The schedule also did not ask about healthcare-specific routes, such as a provider’s complaints office, the Health and Social Services Ombudsman, or the County Governor. The accounts support three things: participants held a general sense of entitlement, could name advocacy organisations when asked, and did not describe acting through any of them. They cannot establish the extent of participants’ knowledge of specific procedures, which was not systematically assessed.

One participant summarised several of these considerations together, giving both an expectation of futility and an account of endurance as the preferable strategy:

> *“I personally feel it is a waste of time going to these institutions considering the long bureaucratic processes. The long process might even emotionally drain you. I have developed thick skin for racial discrimination, and I think that is the best strategy for us Africans.” (Kwasi)*

Participants’ explanations can be organised into five overlapping mechanisms, set out in Table 2. These are interpretive categories, not mutually exclusive or quantified participant types. Table 2 distinguishes those generated during coding from those assembled analytically afterwards.

**Table 2.** Mechanisms of complaint non-use.

| Mechanism | What participants described | Provenance |
| --- | --- | --- |
| Procedural uncertainty† | No reference, in any account, to a specific procedure, form, deadline or office through which a complaint would be made | Analytic category assembled after coding, from the absence of procedural reference across accounts |
| Anticipated futility | An expectation that no useful response would follow, and that the process would be long and bureaucratic | Code generated during coding of the rights theme |
| Relational risk† | Concern that complaining could affect continuing care, expressed as keeping quiet for fear of being denied treatment | Analytic category drawing on material coded under coping |
| Emotional conservation | Avoiding the burden of recounting and pursuing an upsetting event | Code generated during coding of the rights theme |
| Normalisation | Treating discrimination as something to endure rather than an actionable exception, including "developing thick skin" | Code generated during coding of the rights theme |

*† Procedural uncertainty and relational risk are analytic categories assembled after coding; the remaining three mechanisms are codes generated during initial coding of the rights theme (see Provenance column).*

No complaint was described in the interviews. Complaint use was not directly collected, so this cannot establish that no participant had ever complained. From a rights-based perspective, the accounts raise a narrower accountability question. It is whether potential rights-holders perceived mechanisms as sufficiently visible, credible and safe to use while continuing to depend on healthcare.

## Discussion

Ten of the fifteen participants described healthcare encounters they interpreted as discriminatory. All five who described no such encounter had communicated with providers in a shared language. That contrast is unmatched and cannot establish cause, but it locates the study’s central question. The question is whether language accessibility operates only on comprehension, or also on whether patients are recognised as legitimate participants in their own care. These accounts point to the second. They also connect that recognition to what participants did next, which was to continue attending, to avoid care, or to treat themselves informally. Only the first of those is visible in routine utilisation data.

The paper’s principal analytic contribution is integrating candidacy theory, the right-to-health framework and the naming, blaming and claiming tradition into a single account of access and accountability. Together these perspectives suggest a sequence. Language accessibility shapes comprehension and, through it, recognition. Recognition in turn shapes how candidacy is negotiated. Experiences of candidacy then influence whether perceived wrongs become accountability claims. The study therefore proposes that language may bridge accessibility, participation and accountability in the practical realisation of the right to health. In these accounts, communication arrangements and provider conduct shaped how encounters were interpreted. Perceived uncertainty about legitimacy shaped whether candidacy was reasserted or partly withdrawn. Procedural uncertainty, anticipated cost and relational risk shaped whether an interpreted wrong became a complaint. As the conceptual framework states, this is not a causal sequence. It does not imply that every participant moved through every stage, or that one stage caused the next.

That last point speaks to a standing inconsistency in the evidence. Ben et al. [17] found racism associated with poorer healthcare experiences but not consistently with lower healthcare use. These accounts suggest why the two need not move together. Utilisation may reflect need, constrained choice and repeated assertion of candidacy rather than satisfaction, so a count of contacts can record persistence while concealing the harm that produced it. Longitudinal research should test whether clinical need, family composition and viable alternatives modify the relationship between perceived discrimination and service use.

All five contrasting accounts included communication that participants described as adequate in a shared language. We did not match cases on proficiency, residence, education, service type or encounter complexity. This is therefore a hypothesis-generating pattern rather than evidence that language caused the difference. Its importance is conceptual. Formal access to a service may coexist with unequal informational accessibility and participation. We describe this as a tentative linguistic recognition hypothesis. Shared language may facilitate comprehension and, beyond that, recognition, legitimacy and participation within healthcare encounters. Where communication barriers remain unresolved, participants may by contrast experience themselves as peripheral rather than fully recognised. The present study cannot test this proposition causally, but it identifies a direction for comparative research.

Some participants explicitly gave language barriers a social meaning extending beyond technical misunderstanding. They interpreted default use of Norwegian, limited adaptation, or failure to assess comprehension as affecting whether they were recognised as legitimate participants in care. Norwegian equality law is relevant here, because it includes language, skin colour, descent and national origin within ethnicity while not listing race separately [23]. This architecture shows that linguistic disadvantage can intersect with ethnic discrimination. It does not determine that discrimination occurred in these cases.

Three observations support a cautious interpretation of racialisation. First, participants contrasted providers who accommodated communication in English with those who did not, which suggests that institutional response mattered alongside patient proficiency. Second, the reported sick-leave remark assigned a group-based characteristic directly, whilst ambiguous bodily distancing acquired racialised meaning for the participant. Third, some participants explicitly linked a monolingual environment to conditional belonging. These forms of evidence are not equivalent. Together they suggest that language-related barriers may be one pathway through which racialised exclusion is experienced and interpreted.

This formulation also preserves alternative explanations. Communication failure can disadvantage migrants across racialised groups, and inadequate interpretation may arise from workload, misjudged proficiency or resource constraints. Comparable ambiguity has been reported elsewhere in Europe. Sub-Saharan African women using Basque public services described being attended to on professionals’ own communication terms, and questioned whether their treatment reflected racism or misfortune [44]. Future comparative studies should examine whether similar language needs receive different responses across racialised and national-origin groups.

None of the language-related accounts required explicit prejudice to produce unequal consequences. Communication difficulties can arise within a healthcare system organised around a dominant language, even where individual providers intend equitable care. Read this way, language is more than an attribute of individual patients. It operates as an institutional arrangement that shapes accessibility, participation and perceived belonging. Its public-health significance therefore extends beyond translation, to how services anticipate and respond to communicative diversity.

Candidacy clarifies how public-health consequences may arise without complete disengagement. Language and interpreting affected ease of entering and using services, while interpersonal treatment affected perceived legitimacy. Continued attendance represented continued pursuit of care where care remained necessary. Avoidance or self-medication represented reduced engagement with it. These patterns do not establish causal effects. They show why healthcare use alone is an inadequate equity indicator.

Clinical need and limited alternatives were the clearest reasons why continued use could coexist with dissatisfaction. Family responsibility may also have increased the cost of withdrawal in some accounts, and encouraged relationship-preserving behaviour. This complicates descriptions of family solely as a protective resource. The migrant-health literature more commonly treats family and community support as a buffering asset that helps people navigate services [45,46]. Family can support navigation while also creating obligations to remain engaged. Family status was not the basis of a designed comparison and some evidence was second-hand, so this remains a subsidiary hypothesis.

Prompted agreement that discrimination violated rights did not lead to any complaint described in the interviews. This should not be read as proof of procedural ignorance or institutional failure. Participants’ concerns about futility, burden, endurance and ongoing care instead illustrate how accountability is relational. Formally available mechanisms are meaningful only if they are understandable, trusted, independent and experienced as safe.

Candidacy concerned participants’ continuing claim to care. A second and analytically separate question is whether an interpreted wrong could become a claim about the conditions of that care. Read through the naming, blaming and claiming sequence [35], participants’ accounts are consistent with attrition between attribution and claim. They named the treatment as discriminatory. They attributed it to providers or to the system. No participant described converting that attribution into a claim. The schedule did not ask directly whether a participant had ever approached any complaint body, so this cannot establish that they stopped at this specific point rather than simply not being asked. It indicates only that no instance of claiming was volunteered. Perone [36] illustrates that such attrition has been documented elsewhere in healthcare, among long-term care staff who largely did not name residents’ conduct as discrimination at all. That comparison is offered as general illustration only, not as evidence about where attrition occurs here. Perone’s participants were not given a supplied definition of discrimination before describing events, while this study’s were, and staff experiencing residents’ conduct occupy a different structural position from patients experiencing providers’ conduct. What the present accounts show directly is movement from naming to attribution without a described claim. Kwasi’s account locates the block in an assessment of process and cost. Ebe Ano’s locates it in the absence of a channel and the risk of using one. This distinction between claiming care and claiming against care links the candidacy findings to the accountability barriers considered next.

Participants’ expectations of bureaucracy, emotional cost and limited benefit should be taken seriously. The data cannot determine whether those expectations accurately describe each institution. Low Tribunal caseload and compensation figures are contextual indicators, not proof that a complaint would have been futile. How health systems record and respond to complaints of racism is not sufficiently documented for such figures to measure institutional responsiveness [21]. The existence of free, formally low-threshold routes does not by contrast prove practical accessibility. Critical analysis therefore requires linking formal design with lived accessibility. The policy problem is the distance between formal existence and relational complaint capability. A procedure becomes practically usable only when potential complainants can identify it, understand what evidence is expected, get help, view the process as safe for ongoing care, and expect a meaningful response. Healthcare-specific advocates and providers may be better placed than a distant national body to explain options soon after an event. That holds only where complaint handling is independent of clinical decision-making and safeguards against retaliation. The proviso matters, because bringing complaints closer to the service is not automatically an improvement. Kirkland and Hyman [20] found that when discrimination complaints were absorbed into organisational grievance machinery in the United States, they were handled as patient-experience problems and smoothed over with customer-service techniques, which weakened the entitlement rather than realising it. Lenton et al. [22] reached a compatible conclusion from the complainant’s side, arguing that mechanisms which individualise complaints leave the structural conditions untouched. Proximity without independence would reproduce what participants already anticipated.

Future research should follow complaints prospectively, examine case outcomes and compare users who do and do not seek advice. Such work could distinguish procedural complexity, evidentiary difficulty, distrust, lack of information and satisfactory informal resolution. The study contributes to debates about the difference between universal rights on paper and health equity in practice. Norway’s legal and service commitments matter. The practical realisation of the right to health also depends on language accessibility, respectful and acceptable care, meaningful participation, quality communication and accountability. An unexplained or routine clinical practice may acquire exclusionary meaning even where discriminatory intent cannot be established. That patient-level meaning can still affect trust, continuity and public-health outcomes.

### Implications for practice and policy

These implications are offered as hypotheses for policy attention and stakeholder deliberation that the accounts make salient. They are not conclusions this study itself establishes. A sample of fifteen, drawn from one city and skewed towards highly educated, English-speaking residents, cannot by itself warrant system-level prescriptions.

First, treat language accessibility as a quality and equity responsibility rather than a patient-initiated request. Services should proactively assess communication needs. They should document the basis for interpreter decisions. They should provide qualified interpretation where adequate communication cannot otherwise be achieved [28,29].

Second, information accessibility and participation require more than translated written materials. Services should provide information in locally relevant languages and formats. They should use comprehension checks and teach-back. Patients should be able to ask questions and participate meaningfully in decisions. English is not a substitute for qualified interpretation where comprehension is inadequate, or communication is clinically complex.

Third, acceptability and dignity require communication that avoids group-based assumptions and explains unfamiliar practices. Training should address stereotyping, bias, non-verbal communication and the limitations of static cultural-competence models [47–49]. The double-gloving account illustrates how unexplained conduct can acquire harmful meaning even when discriminatory intent cannot be established, and even though plausible non-discriminatory explanations exist, such as glove failure, product substitution or individual habit.

Fourth, complaint and accountability mechanisms should be visible at the point of care, linguistically accessible, and independent of the treating team. Information should distinguish provider complaints, patient-rights and supervisory processes, the Health and Social Services Ombudsman, equality guidance and the Tribunal. Complaints should inform institutional quality improvement rather than be processed solely as isolated dissatisfaction. This is consistent with recommendations elsewhere for a dedicated complaints office addressing racism specifically within inpatient institutions [13,20,22].

Finally, equity monitoring should combine existing country-of-birth measures with ethically governed, voluntary information on self-identified ethnicity, racialisation, language need and patient experience. The purpose is not indiscriminate collection of sensitive data. It is the capacity to identify inequities that healthcare use and country of birth alone may obscure. Rights indicators should assess accessibility, acceptability, participation, quality and accountability, not service contact alone. This monitoring proposal extends beyond what any single account here can support, and would need evaluating against the broader literature on ethnicity data collection before implementation.

### Strengths and limitations

The study provides detailed first-person accounts and includes contrasting cases in which participants reported no discrimination. The shared-language pattern emerged from that comparison. Its analytic value is strengthened by differentiating explicit stereotyping, ambiguous interpersonal conduct and communication failures rather than treating them as equivalent evidence.

The limitations are substantial. The sample was small, selected through purposive and referral methods, based in one city, and skewed towards highly educated, English-speaking residents. It cannot support prevalence claims, nor represent sub-Saharan African immigrants as a homogeneous population. The study did not systematically measure relevant differences in first language, preferred healthcare language, Norwegian proficiency, occupation, family composition or service setting. Eligibility also required at least two prior uses of Norwegian healthcare and a regular general practitioner, so the sample was defined by healthcare use. People who had disengaged from the health service entirely could not appear in it. They are the group for whom avoidance would matter most. Withdrawal is therefore observed here only as partial or episodic reduction among people still attending. Reaching those who no longer attend would require recruitment through community organisations or civil-society networks rather than referral from users.

The shared-language contrast is subject to several conditions wherever it is invoked. We did not match the five contrasting cases with the other ten on length of residence, education, service type or language proficiency, and we did not standardise encounter complexity. We therefore cannot determine whether the pattern reflects shared language, the social advantages associated with it, service context, or some combination. Reports of no discrimination may also reflect differences in expectation, or in willingness to classify an encounter as discriminatory, rather than an objective absence of poor treatment. The contrast identifies a question for comparative research. It does not answer one.

The interview schedule constrains the complaint findings in three specific ways. It named the Ombud and the Tribunal, so awareness of them is not evidence of unprompted knowledge. It asked about advocacy organisations rather than healthcare complaint routes, so silence about the latter is partly a result of the questions. It did not ask directly about use, so non-use is reported from follow-up discussion rather than systematic elicitation. A schedule designed around complaint behaviour would settle what this one can only indicate.

Accounts were retrospective, and telephone interviewing prevented observation of non-verbal conduct. The study contains no provider accounts, clinical documentation or institutional data with which to assess intent, usual practice or alternative explanations. Perceived discrimination is an important outcome in its own right. It should not however be conflated with a legal finding of discrimination. Defining racial discrimination before participants described events may have primed recall and classification, and the five negative cases do not remove that risk.

Evidence for withdrawal and the proposed family-obligation mechanism was limited. It rested on some second-hand material and one developed first-person account of self-medication, in this case informal antibiotic acquisition. Family responsibility is therefore treated as a subsidiary hypothesis rather than a central finding. We did not systematically record participant-level first language, preferred healthcare language, Norwegian proficiency, family composition or encounter complexity, so the negative cases cannot serve as matched comparators. We applied the candidacy framework after coding, and the study was not designed to test the integrated sequence prospectively. Future research should use longitudinal, multi-site and comparative designs, and pair patient accounts with provider, complaint and organisational data.

## Conclusions

Universal entitlement did not deliver equal access in these accounts. Language accessibility operated as more than a technical matter of comprehension. Participants read the adequacy of communication as a signal of whether they were recognised as legitimate participants in their care. Where that recognition was absent, some continued to attend because they had to. Others withdrew or treated themselves.

Two consequences follow. Healthcare use is an inadequate measure of equitable access. Attendance can record clinical need or the absence of alternatives rather than acceptable care, so monitoring that counts contacts will not detect the harm described here. Accountability mechanisms that racialised patients do not use also generate institutional records showing no problem. That is not the same as an absence of one. No participant described converting an interpreted wrong into a claim, although the schedule did not ask directly.

Comparative work should now test whether similar language needs receive different responses across racialised and national-origin groups. Such designs should match cases on proficiency, length of residence and service setting rather than relying on contrast alone. Complaints should also be followed prospectively and paired with institutional data on how healthcare organisations record and respond to allegations of racism. That would establish whether attrition occurs where these accounts suggest it does. Both are needed before language accessibility can be treated as a measured determinant of equitable care rather than a plausible one.

## Supporting information

Correct Ethical Approval document

## Data Availability

The qualitative transcripts contain potentially identifying information from a small community and are not publicly available. De-identified excerpts beyond those reported may be considered by the corresponding author on reasonable request, subject to the original ethics approval and applicable data-protection requirements.

## Acknowledgments

The authors thank the participants for sharing their experiences.

## Author Contributions

Peter Kofi Taadi: Conceptualization, Methodology, Investigation, Formal analysis, Writing - original draft. Bill Derman: Supervision, Writing - review & editing.

## Supporting information

S1 File. COREQ checklist. Consolidated Criteria for Reporting Qualitative Research, completed against each of the 32 items with section-level pointers to the main text.

S2 File. Coding tree. Themes, sub-themes and illustrative codes generated during reflexive thematic analysis, with a note distinguishing codes generated during coding from analytic categories assembled afterwards.

S3 File. Interview guide. Semi-structured interview schedule used for data collection, including both branch versions and a note on the screening question’s compound wording.

## List of abbreviations

COREQ: Consolidated Criteria for Reporting Qualitative Research
GP: general practitioner
ICERD: International Convention on the Elimination of All Forms of Racial Discrimination
NSD: Norwegian Centre for Research Data
OHCHR: Office of the United Nations High Commissioner for Human Rights
WHO: World Health Organization.

