## Supplementary material for "Language, perceived discrimination and barriers to complaint use: sub-Saharan African immigrants’ healthcare experiences in Oslo, a qualitative interview study": Correct Ethical Approval document

Sikt

### Assessment of processing of personal data

| Reference number | Assessment type | Date |
| --- | --- | --- |
| 186966 | Standard | 19.06.2020 |

**Title**
Racial discrimination among sub Saharan Africans using health care services in Norway

**Institution responsible for the project**
Norges miljø- og biovitenskapelige universitet (NMBU) / Fakultet for landskap og samfunn / Institutt for internasjonale miljø- og utviklingsstudier

**Project leader**
William Derman

**Student**
PETER KOFI TAADI

**Academic level**
Master's

**Processing period**
03.08.2020 – 30.05.2021

**Categories of personal data**
General
- Name
- Date of birth
- Voice on audio recordings

Special
- Ethnicity

**Legal basis**
Consent, cf. GDPR Article 6(1)(a)
Explicit consent, cf. GDPR Article 9(2)(a)

Description of the processing

**Research purpose and justified need for personal data**
The purpose of the study is to explore the racial discrimination experiences of immigrants living in Norway when they make use of healthcare services. Qualitative study design shall be used to collect data from participants after obtaining their consent. Among other things, the study will seek to find out if they experience racial discrimination and in what forms if they do. It shall also explore how they cope with such experiences and their perceived effect on their health. The study shall not collect data on specific disease conditions of participants.

1. it will help us to identify the different means by which the different genders and age group though from the same race experience discrimination.

No

**Total number of data subjects in the project**
1-99

#### Sample 1

sub Saharan Africans in Norway

initial contact will be made through my network for example , the African students union, Africans association in Norway etc.

Age group: 18 - 120

**Data collection/method**
- Personal interview

#### Overall assessment

> We have assessed that the processing of personal data will meet requirements in data protection legislation. We carry out assessments on behalf of the data controller. You must follow the data controller's guidelines for information security and any conditions included in the assessment.

Our assessment is that the processing of personal data in this project will comply with data protection legislation, so long as it is carried out in accordance with what is documented in the Notification Form and attachments, dated 19.20.2020, as well as in correspondence with NSD. Everything is in place for the processing to begin.

SHARING THE PROJECT WITH THE RESPONSIBLE FOR THE PROJECT It is mandatory for students to share the project with their supervisor. You can do this by clicking "Share project" in the upper left corner. By selecting "Invite user", you can add the e-mail address of the people you want to share with, as well as administer access ("Can edit" gives access to making changes in the form), make sure that the e-mail address is correctly spelled. Note that they will also have access to the "Data management plan" (if you have one) and to see the dialogue with us, however they do not have the option to send messages.

NOTIFY CHANGES If you intend to make changes to the processing of personal data in this project it may be necessary to notify NSD. This is done by updating the Notification Form. On our website we explain which changes must be notified. Wait until you receive an answer from us before you carry out the changes.

TYPE OF DATA AND DURATION The project will be processing special categories of personal data about racial or ethnic origin , and general categories of personal data, until 30.05.2021.

LEGAL BASIS The project will gain consent from data subjects to process their personal data. We find that consent will meet the necessary requirements under art. 4 (11) and 7, in that it will be a freely given, specific, informed and unambiguous statement or action, which will be documented and can be withdrawn.

The legal basis for processing special categories of personal data is therefore explicit consent given by the data subject, cf. the General Data Protection Regulation art. 6.1 a), cf. art. 9.2 a), cf. the Personal Data Act § 10, cf. § 9 (2).

PRINCIPLES RELATING TO PROCESSING PERSONAL DATA NSD finds that the planned processing of personal data will be in accordance with the principles under the General Data Protection Regulation regarding:

- lawfulness, fairness and transparency (art. 5.1 a), in that data subjects will receive sufficient information about the processing and will give their consent
- purpose limitation (art. 5.1 b), in that personal data will be collected for specified, explicit and legitimate purposes, and will not be processed for new, incompatible purposes
- data minimisation (art. 5.1 c), in that only personal data which are adequate, relevant and necessary for the purpose of the project will be processed
- storage limitation (art. 5.1 e), in that personal data will not be stored for longer than is necessary to fulfil the project’s purpose

THE RIGHTS OF DATA SUBJECTS Data subjects will have the following rights in this project: transparency (art. 12), information (art. 13), access (art. 15), rectification (art. 16), erasure (art. 17), restriction of processing (art. 18), notification (art. 19), data portability (art. 20).

These rights apply so long as the data subject can be identified in the collected data.

NSD finds that the information that will be given to data subjects about the processing of their personal data will meet the legal requirements for form and content, cf. art. 12.1 and art. 13.

We remind you that if a data subject contacts you about their rights, the data controller has a duty to reply within a month.

FOLLOW YOUR INSTITUTION’S GUIDELINES NSD presupposes that the project will meet the requirements of accuracy (art. 5.1 d), integrity and confidentiality (art. 5.1 f) and security (art. 32) when processing personal data.

To ensure that these requirements are met you must follow your institution’s internal guidelines and/or consult with your institution (i.e. the institution responsible for the project).

FOLLOW-UP OF THE PROJECT NSD will follow up the progress of the project at the planned end date in order to determine whether the processing of personal data has been concluded.

Good luck with the project!

Contact person at NSD: Henriette N. Munthe-Kaas Data Protection Services for Research: +47 55 58 21 17 (press 1)
